# Changes in vaping trends since the announcement of an impending ban on disposable vapes: a population study in Great Britain

**DOI:** 10.1101/2024.12.19.24319341

**Authors:** Sarah E. Jackson, Lion Shahab, Harry Tattan-Birch, Vera Buss, Jamie Brown

**Affiliations:** Department of Behavioural Science and Health, University College London, London, UK; SPECTRUM Consortium, UK

**Keywords:** vaping, e-cigarettes, disposable vapes, Tobacco and Vapes Bill, disposable vape ban, population trends

## Abstract

**Background/Aim:** There has been a rapid rise in vaping prevalence among youth and young adults in Great Britain since disposable vapes started to become popular in 2021. In January 2024, the government announced plans to introduce a number of new vaping policies, including a ban on disposable vapes, to tackle youth vaping. This study examined whether trends in current vaping and use of disposable vapes have changed since this announcement.

**Design, setting, and participants:** Segmented regression analysis of data collected monthly between January 2022 and November 2024 as part of the Smoking Toolkit Study; a representative household survey in Great Britain. We ran generalised additive models using data from all participants aged ≥16y (*n*=83,764; ‘adults’) and from a subset aged 16-24y (*n*=8,846; ‘young adults’).

**Main outcome measures:** Changes in trends in (i) the prevalence of current vaping and (ii) the proportion of vapers mainly using disposable devices.

**Results:** Before January 2024, vaping prevalence was increasing by 24.5% per year (RR_trend_=1.245 [95%CI 1.193-1.299]) and use of disposable vapes was increasing by 18.0% per year (RR_trend_=1.180 [95%CI 1.106-1.258]). These trends changed after the new policy measures were announced (RR_Δtrend_=0.718 [0.623-0.827] and RR_Δtrend_=0.558 [0.460-0.677], respectively). Instead of increasing, there was an uncertain decrease in vaping prevalence from 13.7% [12.7-14.7%] in January 2024 to 12.4% [11.4-13.6%] in November 2024) and a substantial decline in the proportion of vapers mainly using disposables from 43.7% [40.3-47.3%] to 30.8% [27.6-34.4%]. Similar changes were observed among young adults (vaping prevalence: RR_Δtrend_=0.647 [0.495-0.845]; use of disposable vapes: RR_Δtrend_=0.526 [0.400-0.693]), with an uncertain decline in vaping prevalence from 27.7% [24.5-31.3%] to 23.6% [20.2-27.6%] between January and November 2024 and the proportion of vapers mainly using disposables falling from 63.4% [59.0-68.2%] to 37.8% [32.3-44.2%].

**Conclusions:** Following the announcement of an impending ban on disposable vapes and other potential vaping policies, recent increases in vaping prevalence in Great Britain stalled – or may have even reversed – including among young adults. In addition, there was a shift away from using disposable vapes, with people increasingly opting to use devices that can be refilled and recharged.

## Introduction

The prevalence of e-cigarette use (‘vaping’) among youth and young adults in Great Britain has risen rapidly since 2021.^1–4^ This increase appears to have been driven by the introduction of new disposable vapes, which discarded after the battery or e-liquid runs out.^1–3,5,6^ These products are cheap and easy to use, have sleek design and colourful branding, and are widely available from a range of retail outlets.^7–9^ As of 2023, most young vapers were using disposables,^3,4^ as were around half of adults who had been vaping for six months or more.^4^ Concerns over their appeal to youth and their environmental impact^10^ led to calls for an outright ban on disposable vapes.^11–13^

In October 2023, the UK Government published a command paper outlining a range of measures it was considering to reduce youth vaping, including restricting the sale of disposable vapes.^14^ Following a six-week consultation, the Prime Minister announced in January 2024 that disposable vapes would be banned in the UK as part of a package of measures\ ‘to tackle the rise in youth vaping and protect children’s health’.^15^ In addition to reducing youth vaping, another important rationale for banning disposables was that it would have benefits for the environment.^15^ This announcement attracted widespread media coverage.^e.g., 16^

New legislation to ban the sale of disposable vapes in England was developed under the Environmental Protection Act 1990 and will come into effect from 1 June 2025.^17,18^ The Scottish and Welsh Governments have also confirmed that they will align with England and introduce a ban on disposables on the same date.^19,20^ Other measures to tackle youth vaping are currently progressing through parliament as part of the Tobacco and Vapes Bill.^21^ These include powers to restrict the way e-cigarettes are formulated (e.g., the flavours they can contain), branded (e.g., the way they are packaged), marketed, and sold (e.g., the way they are displayed in shops), all of which will be subject to consultation and secondary legislation.

However, the vaping landscape evolves rapidly and changes in behaviour are often seen before legislation comes into effect.^22^ E-cigarette manufacturers responded to news of the ban by starting to sell ‘disposable-like’ products: reusable versions of their popular disposable models. Notifications to the Medicines and Healthcare products Regulatory Agency for disposable products quickly declined following the announcement of the ban, while notifications for products with similar branding and features to those in disposable category but that were newly rechargeable or refillable increased.^23^ This may have prompted some vapers to transition away from disposable devices. Other vapers may have sought out reusable options after hearing about the impending ban. News coverage of the adverse impacts of disposable vapes on the environment may have increased the salience of these concerns for some users.

It is important to have up-to-date information to inform policy decisions. This study aimed to establish whether there have been changes in e-cigarette use in the months since the government announced its intention to introduce new vaping policies. Specifically, we aimed to examine changes in monthly trends in (i) vaping prevalence and (ii) the proportion of vapers who report mainly using disposable devices, among people in Great Britain aged ≥16 years (‘adults’) and specifically among those aged 16-24 years (‘young adults’).

## Methods

### Design

Data were drawn from the Smoking Toolkit Study, an ongoing monthly cross-sectional survey of a representative sample of adults (≥16 years) in Great Britain.^24,25^ The study uses a hybrid of random probability and simple quota sampling to select a new sample of approximately 2,450 adults each month. Data are collected through telephone interviews. Comparisons with other national surveys and sales data indicate the survey achieves nationally representative estimates of key sociodemographic and nicotine use variables.^24,26^

The present analyses used data from respondents in the period from January 2022 (around six months after disposable vapes started to become popular^1^ and two years before the government’s announcement of new potential vaping policies^15^) to November 2024 (the most recent data available at the time of analysis). Data on the main device type used were not collected in England in May, June, or August 2022; these waves were therefore excluded from analyses of trends in the use of disposable vapes.

### Measures

Vaping status was assessed within several questions asking about use of a range of nicotine products. Participants who reported current smoking were asked ‘Do you regularly use any of the following in situations when you are not allowed to smoke?’ and those who reported cutting down ‘Which, if any, of the following are you currently using to help you cut down the amount you smoke?’; those who currently smoked or who had quit in the past year were asked ‘Can I check, are you using any of the following either to help you stop smoking, to help you cut down or for any other reason at all?’; and non-smokers were asked ‘Can I check, are you using any of the following?’. Those who reported using an e-cigarette in response to any of these questions were considered current vapers, else they were considered non-vapers.

Main device type used was assessed by asking vapers: ‘Which of the following do you mainly use…?’ (a) a disposable e-cigarette or vaping device (non-rechargeable); (b) an e-cigarette or vaping device that uses replaceable pre-filled cartridges (rechargeable); (c) an e-cigarette or vaping device with a tank that you refill with liquids (rechargeable); or (d) a modular system that you refill with liquids (you use your own combination of separate devices: batteries, atomizers, etc.). We categorised device types as disposable (response *a*) or reusable (responses *b-d*). Participants could also respond that they did not know; these cases were treated as missing.

### Statistical analysis

These analyses were not pre-registered and should be considered exploratory. Data were analysed in R v.4.2.2. The Smoking Toolkit Study uses raking to create survey weights that match the sample to the population in Great Britain.^25^ The following analyses were done on weighted data. Missing cases were excluded on a per-analysis basis.

We used segmented regression to assess changes in monthly trends in current vaping and mainly using disposable vapes in Great Britain following the government’s announcement of new policy measures to reduce youth vaping. We used log-binomial generalised additive models (GAMs, using the *mgcv* package in R) to model trends before the announcement (underlying secular trend; coded January 2022 = 1 through November 2024 = 35) and the change in the trend (slope) after the announcement (coded 0 up to January 2024 and 1…*n* from February 2024 onwards, where *n* was the number of waves after the announcement). A step-level change was not included as this was deemed implausible. To adjust for seasonality (month-of-year effects), we also included a variable reflecting the calendar month coded from January=1 to December=12; this variable was modelled using a smoothing term with cyclic cubic splines specified. We assumed a linear trend in log-prevalence before the announcement (i.e., the proportional change in prevalence month-on-month would be stable from January 2022 to January 2024). Because of the relatively short length of the time-series, we expected negligible differences between log-linear and linear trends. To explore changes among young adults specifically, we repeated the analyses restricting the sample to participants aged 16-24y. We used predicted estimates (using the *predict* function) to plot modelled trends in each outcome alongside unmodelled data points and to estimate the proportion of vapers likely to be mainly using disposables by the time the ban on disposable vapes is implemented in June 2025 (calculated by using the model to predict estimates over an extended time period).

## Results

A total of 83,764 participants were surveyed between January 2022 and November 2024 (mean [SD] age = 48.1 [18.9]; 50.6% women), of whom 8,846 were aged 16-24y (mean [SD] age = 20.7 [2.4]; 47.6% women). There were 7,424 vapers surveyed in waves in which the main device type used was assessed; 4.9% (365/7,424) did not know the main device type they used and were excluded from analyses of trends in use of disposable vapes.

Following the announcement in January 2024 of an impending ban on disposable vapes and other potential vaping restrictions, there were notable changes in trends in e-cigarette use in Great Britain (**Table 1**).

**Table 1.** GAM results: changes in trends in e-cigarette use since the announcement of an impending ban on disposable vapes in January 2024.

| | Adults ($\geq 16y$) | | | Young adults (16-24y) | | |
| --- | --- | --- | --- | --- | --- | --- |
|  | RR | 95% CI | p | RR | 95% CI | p |
| **Current vaping<sup>1</sup>** |  |  |  |  |  |  |
| Trend (year) | 1.245 | 1.193-1.299 | <0.001 | 1.277 | 1.180-1.382 | <0.001 |
| ΔTrend (year) | 0.718 | 0.623-0.827 | <0.001 | 0.647 | 0.495-0.845 | 0.001 |
| **Mainly uses disposable vapes<sup>2</sup>** |  |  |  |  |  |  |
| Trend (year) | 1.180 | 1.106-1.258 | <0.001 | 1.021 | 0.944-1.105 | 0.599 |
| ΔTrend (year) | 0.558 | 0.460-0.677 | <0.001 | 0.526 | 0.400-0.693 | <0.001 |

RR, risk ratio; CI, confidence interval; GAM, generalised additive model.
Results shown are segmented regression analyses of data collected from January 2022 to November 2024.
<sup>1</sup> Among all respondents (*unweighted n: 83,764 adults and 8,846 young adults*).
<sup>2</sup> Among respondents who reported current vaping in waves that assessed the main device type used (*unweighted n: 7,059 adults and 1,594 young adults*).
Models are adjusted for seasonality. Note: the model for trends in use of disposable vapes among adults ( $\geq 16y$ ) had problems with convergence, so seasonality was modelled bi-monthly rather than monthly.
Trend is the underlying secular trend. $\Delta$ Trend is the change in trend (slope) following the announcement of an impending ban on disposable vapes and other potential vaping restrictions.

Between January 2022 and January 2024, the prevalence of current vaping increased by 24.5% per year among adults (RR_trend_=1.245) from 8.8% [8.1-9.6%] in January 2022 to 13.7% [12.7-14.7%] in January 2024 and by 29.1% per year among young adults (RR_trend_=1.291) from 17.0% [14.8-19.5%] to 27.7% [24.5-31.3%] (**Figure 1A**). The proportion of vapers who mainly used disposable devices increased by 18.0% per year among adult vapers (RR_trend_=1.180) from 31.4% [28.0-35.2%] to 43.7% [40.3-47.3%] and was approximately stable among young adult vapers (RR_trend_=1.021) at a majority of 62.1% on average [58.2-66.3%] (**Figure 1B**).

**Figure 1.**
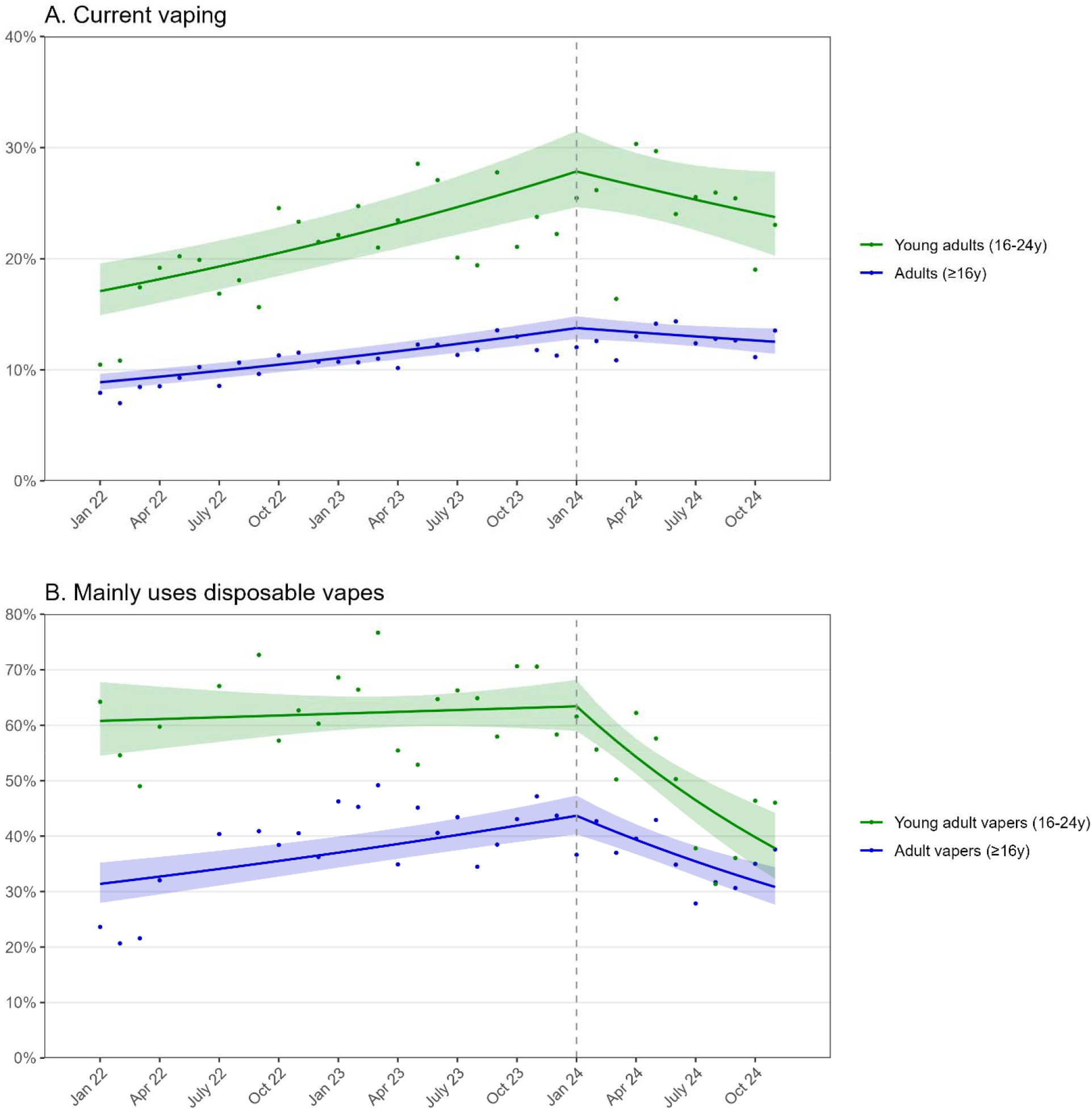
Trends in current vaping and use of disposable vapes, January 2022 to November 2024. Panels show trends in the prevalence of (A) current vaping among adults and young adults, and (B) predominant use of disposable vapes among adult and young adult vapers. The vertical dashed line indicates the timing of the announcement in January 2024 of an impending ban on disposable vapes and other potential vaping restrictions. Points represent unmodelled weighted prevalence by month. Lines represent modelled weighted prevalence over the study period, assuming log-linear trends pre- and post-announcement and adjusting for seasonality. Shaded bands represent 95% confidence intervals.

After the new policy measures were announced in January 2024, the trend in current vaping prevalence among all adults changed (RR_Δtrend_=0.718, *p*<0.001; **Figure 1A**): prevalence was no longer increasing and there was instead an uncertain decline of -10.6% per year [RR_trend_=1.245 x RR_Δtrend_=0.718 = 0.894] from 13.7% [12.7-14.7%] in January to 12.4% [11.4-13.6%] in November 2024. Similarly, the trend in current vaping prevalence among young adults changed (RR_Δtrend_=0.647, *p*=0.001) resulting in an uncertain decline of -17.4% per year [RR_trend_=1.277 x RR_Δtrend_=0.647 = 0.826]) from 27.7% [24.5-31.3%] to 23.6% [20.2-27.6%].

There was also a substantial downward change in the trends in the use of disposable vapes (adult vapers: RR_Δtrend_=0.558, *p*<0.001; young adult vapers: RR_Δtrend_=0.526, *p*<0.001; **Figure 1B**). The proportion of vapers mainly using disposable devices declined by 34.2% per year (RR_trend_=1.180 x RR_Δtrend_=0.558 = 0.658) among adult vapers (from 43.7% [40.3-47.3%] in January 2024 to 30.8% [27.6-34.4%] in November 2024) and by 46.2% per year (RR_trend_=1.021 x RR_Δtrend_=0.526 = 0.537) among young adult vapers (from 63.4% [59.0-68.2%] to 37.8% [32.3-44.2%]). If current trends continue at the same rate, we project that an estimated 24.2% [20.0-29.1%] of adult vapers and 26.3% [19.8-35.0%] of young adult vapers would be mainly using disposable vapes by the time the ban on disposables is implemented in June 2025.

## Discussion

Since the government announced its intention to ban disposable vapes and to introduce other potential vaping restrictions in January 2024, there have been two important changes in vaping trends in Great Britain. First, people have increasingly opted to mainly use reusable rather than disposable vapes. Based on current trends, we estimate that the proportions of adult and young adult vapers mainly using disposable vapes will have roughly halved between the announcement of the ban in January 2024 and its implementation in June 2025. Second, recent increases in vaping prevalence have stalled – and may even be reversing – including among young adults.

Our findings provide some reassurance that vaping rates are no longer continuing to rise rapidly. Therefore, the stricter policy options under the Tobacco and Vapes Bill are not necessary to halt the rapid rise in vaping, but some restrictions may be needed to bring the high rates of vaping that persist down further. While vapers shifting away from disposable products is likely to have a positive impact on reducing environmental harms,^10^ it does mean that a ban on disposable vapes is unlikely to have a large impact on reducing vaping prevalence. Other policy measures are likely to be needed to make vaping less appealing to young people. It will be important to weigh the need for tighter regulation of vaping to reduce appeal to young people against any unintended consequences for smokers (who may benefit from using e-cigarettes to stop smoking^27^) and for ex-smokers who vape (for whom vaping provides a less harmful alternative to smoking^28^).

As our analysis demonstrates, Great Britain has detailed and regular monitoring of vaping and smoking outcomes which can provide real-time surveillance and allow policy decisions to be informed by the latest available evidence. A limitation is that the survey only captures the main device type used by vapers; in reality, numbers using disposable vapes will be higher given that some people will use both disposable and reusable vapes. Nonetheless, our data provide useful insights into changes in vaping prevalence and product choice in a rapidly evolving policy landscape.

## Data Availability

Data are available on Open Science Framework (https://osf.io/fsudk/).

## Declarations

### Ethics approval

Ethical approval for the STS was granted originally by the UCL Ethics Committee (ID 0498/001). The data are not collected by UCL and are anonymised when received by UCL.

